# Nationwide estimates of SARS-CoV-2 infection fatality rates and numbers needed to vaccinate for SARS-CoV-2 vaccines in 2024 in Austria

**DOI:** 10.1101/2025.08.20.25334029

**Authors:** Uwe Riedmann, Martin Sprenger, John PA Ioannidis, Stefan Pilz

**Affiliations:** Department of Internal Medicine, Division of Endocrinology and Diabetology, Medical University of Graz, 8036 Graz, Austria; Institute of Social Medicine and Epidemiology, Medical University of Graz, 8036 Graz, Austria; Departments of Medicine, Epidemiology and Population Health, and Biomedical Data Science, and Meta-Research Innovation Center at Stanford (METRICS), Stanford University, Stanford, CA, USA

**Author notes:** Corresponding Author: Stefan Pilz, MD, PhD Medical University of Graz, Department of Internal Medicine, Division of Endocrinology and Diabetology Auenbruggerplatz 15, 8036 Graz, Austria.

**Keywords:** COVID-19, Infection fatality rate, vaccination, Number needed to vaccinate, Life-year saved

## Abstract

**Background:** Post-pandemic years are characterized by widespread previous population immunisation against COVID-19. Whether and for whom SARS-CoV-2 vaccinations are still justified is unclear. We use nationwide estimates of IFR and literature derived estimates of vaccine effectiveness (VE) to calculate numbers needed to vaccinate to prevent one COVID-19 death (NNV) and for one life-year saved (LYS) in Austria in 2024.

**Methods:** In this retrospective analysis we calculate SARS-CoV-2 IFR during 2024 in Austria according to previously published wastewater-based infection estimates and available mortality data. Using literature derived VE estimates we calculate NNV to prevent one COVID-19 death and for one LYS in strata according to age groups, nursing home residency and vaccination in 2024. We repeat analyses with sensitivity range values of parameters.

**Results:** In 2024, total IFR was 0.048%. NNV (LYS) in the age groups 0-19, 20-39, 40-59, 60-74 and 75-84 years were very high: e.g. 5,497,526 (151,570), 2,432,498 (92,614), 415,714 (24,777), 35,925 (3,748), and 4,882 (1,009), respectively, in community dwellers. In the 85+ years age group, IFRs of unvaccinated/vaccinated were 0.91%/0.77% for community dwellers, and 1.22%/1.04% for nursing home residents. The 85+ year age group had NNV estimates of 1,215 and 907 (LYS: 525 and 1,896) in community dweller and nursing home residents, respectively. Sensitivity analyses yielded LYS<1,000 only under some favourable assumptions in the 75-84 and 85+ years old age strata.

**Conclusions:** In 2024 SARS-CoV-2 IFR was low and NNV and LYS of COVID-19 vaccinations correspondingly non-favourably high, even for very old individuals.

## INTRODUCTION

The unprecedentedly rapid development of COVID-19 vaccines is credited with potentially saving several millions of lives during the pandemic.^1^ In the current endemic phase, decreasing infection fatality rate (IFR) and relatively low or uncertain booster effectiveness in previously infected individuals raise questions about whether continued vaccination is still justifiable.^2–5^ Knowledge gaps regarding this issue are reflected by huge differences in country-specific COVID-19 vaccine policies.^6–8^

Early randomized controlled trials (RCTs) showed high relative SARS-CoV-2 vaccine efficacy, but in the endemic phase virtually everyone has previously been infected and the majority of the population has also been vaccinated, providing high protection against severe COVID-19 outcomes.^2,5,9–11^ In this environment, studies on VE have been less conclusive, and were often performed in especially high-risk groups not representative of the general population, e.g. immunocompromised individuals.^3,12–14^ While our previous study^12^ indicated low booster VE against infection (17%) and no significant VE against COVID-19 deaths in Austria, a recent test-negative case-control study found 48% VE against COVID-19–associated critical illness.^15^ However, these latter VE estimates incorporated periods where no vaccine effect is expected (i.e. within two weeks of vaccination), which may indicate the presence of a healthy vaccinee bias, i.e. vaccinated individuals are healthier than the unvaccinated controls.^16–18^ Crucially, even if the relative effectiveness of vaccines was very high, low disease severity may still indicate that further vaccinations are not justifiable from a cost-effectiveness perspective, and that COVID-19 vaccination programs may need to be tailored to specific groups of patients.^19,20^

Up to date estimates that are representative of the SARS-CoV-2 infection rate and the severity of COVID-19 in the general population (e.g., IFR), and current estimates of VE of COVID-19 vaccines are required to inform current public health policies regarding COVID-19, but such data are sparse.^4,21^ As a consequence, there are widely varying COVID-19 vaccination recommendations across countries (e.g., in 2024, vaccination was recommended for individuals 12 years and older in Austria and 65 years and older in Denmark).^6,7,20^ Importantly, as the risk and vaccine impact differs based on demographics (especially age and nursing home residency), individual estimates for different age and nursing home strata are essential.^4,22^

In this nationwide retrospective analysis, we estimated SARS-CoV-2 infections, the severity of COVID-19 (via IFR), and VE in 2024 in Austria to calculate numbers needed to vaccinate (NNV) to prevent one COVID-19 death, and for one life-year saved (LYS). We used previously published wastewater based estimates of SARS-CoV-2 infections to calculate IFRs for different strata based on age group, vaccination status (i.e., vaccinated in 2024 or not) and nursing home residency.^5,23^ We achieved this by allocating infections and deaths to different strata based on conditional VE estimates and infection probability assumptions.^24^ We additionally performed sensitivity analyses with different parameter values.

## METHODS

### Study design and analysis

We conducted a retrospective estimation of strata-specific SARS-CoV-2 IFRs, and COVID-19 vaccine NNV and LYS in Austria in 2024. Strata were based on age groups, nursing home residency and whether the individuals were vaccinated in 2024. National estimates of SARS-CoV-2 infections in Austria were derived from wastewater data as published previously by our group.^23^ Data (by age group) on COVID-19 mortality, nursing home population, life tables and vaccination were derived from publicly available sources.^25–28^ To allocate infections and deaths to the respective strata, we used literature-based estimates for VE against infection (VEI) and conditional VE against death, given an infection (VED),^12,15^ as well as reported age group dependent infection probabilities^29^, published nursing home residency dependent infection probabilities^30^ and published age group-specific proportions of COVID-19 deaths between nursing home residents and the community dwellers in Austria.^4^ Life tables were based on the most recent available (2023) estimates.^27^

We followed the Strengthening the Reporting of Observational Studies in Epidemiology (STROBE) checklist (Table S1). The study was approved by the ethics committee at the Medical University of Graz (no. 33-144 ex 20/21). The statistical analyses were conducted using R (version 4.4.2).^31^

### Assumptions

We assumed that every individual was infected at least once before January 2024, thus VED and VEI are based on hybrid immunity estimates.^32^ We also assumed that vaccination probability is the same in nursing home residents and community dwellers within an age group. It is important to note that throughout the paper the term “vaccinated” and “unvaccinated” refers to individuals that did/didn’t get vaccinated in 2024, irrespective of previous vaccination status. That is because we don’t differentiate between primary or booster doses, as an available review does not indicate clear VEI differences.^3^ While currently there are no robust findings on VED comparing hybrid immune that only had a primary vaccination, and those that also had booster doses, the same review found no difference in “hospital admission or severe disease” between the groups. ^3^

### Calculations

The foundation for further calculations was set by the allocation of SARS-CoV-2 infections and COVID-19 deaths to the strata. In short, we separated infections into vaccinated and unvaccinated based on VEI and then allocated them into different age group and nursing home residency strata based on population size as well as literature-based age group and nursing home status dependent infection probabilities distribution (age group infection probability = AIP; nursing home infection probability = NIP). Estimated infected individuals were then used to further allocate COVID-19 deaths to vaccinated and unvaccinated groups (within age groups) based on VED estimates and group population size. Nursing home allocation of deaths was based on group population size and COVID-19 mortality proportions between nursing home residents and community dwellers in different age groups, as reported for the Austrian pandemic.^4^ The allocation process is described in depth in the supplements.

The allocated deaths were divided by the allocated infections to calculate the IFR. From the allocated deaths, we additionally calculate the absolute risk reduction (ARR) of vaccines for every stratum(i) of individuals not vaccinated in 2024^21^:

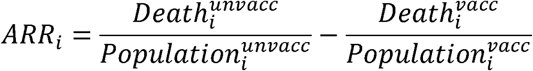

We could then derive the number needed to treat (NNV) to avert one death for every stratum:

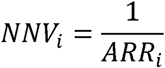

Similarly, number needed to vaccinate to save one life-year (LYS) was calculated proportional to NNV by the weighted average life expectancy (LE) within the respective stratum:

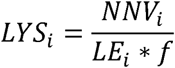

It is known that individuals who died due to COVID-19 were more debilitated than other individuals in the same strata.^1^ We used a scalar factor f to account for a reduced LE of individuals plausibly saved by a vaccination, akin to a previous publication.^1^

### Values used and sensitivity analyses

Strata were based on age (in years) groups (0-19, 20-39, 40-59, 60-74, 75-84, 85+), nursing home residency (for 60-74, 75-84 and 85+ age groups only) and received vaccination in 2024 (over 440,000) in the entire Austrian population (9,197,213 by January 1, 2024).^25–27,33^

### Infections

SARS-CoV-2 infections were estimated at 2.5 million in 2024.^23^ Sensitivity range was set to 2 million to 3.5 million. The upper end was chosen relatively high as there are studies indicating a decrease in viral shedding in the previously infected population, which would lead to underestimation of infections in the wastewater based estimates.^23,34,35^

### Age and nursing home dependent infection rates

We defined relative rates for how likely an age group is to be infected, compared to other age groups. A review of early seroprevalence data suggests no clear trend of infection probability between children, non-elderly adults and the elderly.^36^ The relative infection distribution (age group infection probability; AIP) were thus set to 1 for all age group (equal chance of getting infected). For sensitivity analyses conditions we set (a) values of the three older groups (60-74, 75-84, 85+) to 0.5 (“low” condition) or (b) to 2 (“high” condition) respectively. Some studies have also shown that the probability of infection in nursing homes may have been much higher than for community dwellers early in the pandemic.^30,37^ However, heighted attention was given to nursing home subsequently and, to our knowledge, no reliable data exists for post-pandemic evaluation. In the main analysis, we used equal probability to get infected in 2024 irrespective of nursing home residency (nursing home infection probability = 1; NIP).^36^ We used high variation in the sensitivity range to account for the high uncertainty: (“low”: 0.5 times the infection rate in nursing home residents; “high”: 4 times higher infection rates in nursing home residents).

### Vaccine effectiveness

We used the estimate of our previous nationwide study in Austria as the source for the VEI estimate VEI= 20% (sensitivity range: 5% - 40%).^12^ Estimates on VED comparing natural immune and hybrid immune are scarce. A recent study from the US indicates a 48% VE against COVID-19–associated critical illness.^3,15^ This estimate is, however, driven by high early protection against infection (VEI), as the findings show no significant protection after 6 months.^15^ This is in line with the fast waning found in VEI, and indicates low VED as previous studies show much slower waning in protection against severe cases.^11,12^ Additionally, recent studies show healthy vaccinee effects in epidemiological analyses of COVID-19 VE.^16–18^ Thus, we assumed a relatively low VED of 15% (sensitivity range: 5% – 35%). This combination of conditional VEI and VED leads to an estimated VE of 32% (Figure S1), calculated as such^24^:

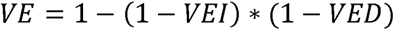

Possible underestimation of VE was addressed by the combination of high sensitivity range value of VEI (40%) and VED (35%) which corresponds to a VE of 61% (Figure S1).

### Life expectancy

LE of every age was weighted by population size to calculate age group LE values (community dwelling) that can be found in Table 1 and are required to calculate LYS. The average LE of nursing home residents is vastly lower than for the community dwelling populations.^38^ For nursing home resident in the 75-84 year age group we used an LE estimate of 2 years.^1^ Community dweller LE in the 75-84 year age group was 4.84 times higher than nursing home resident LE. We used the same ratio to estimate 60-74 and 85+ age group nursing home resident LE, based on their respective community dwelling LE (Table 1). For factor f we chose 0.5 (sensitivity 0.25 – 0.8), as suggested previously.^1^ Smaller values assume that those who die are in worse health than the respective same-stratum general population.

**Table 1:** Descriptive statistics overall and of different age groups in Austria in 2024.

|  | Population<br>(n) | Nursing<br>home<br>residents<br>(n) | Vaccinated<br>in 2024 (n)* | COVID-19<br>deaths (n) | LE<br>community<br>dwellers<br>(years) | LE nursing<br>home<br>residents<br>(years)* |
| --- | --- | --- | --- | --- | --- | --- |
| Total | 9,167,923 | 66,340 | 441,452 | 1,212 | - | - |
| 0-19 year old | 1,761,530 | 0 | 8,708 | 1 | 72.54 | - |
| 20-39 year old | 2,347,974 | 0 | 40,568 | 3 | 52.53 | - |
| 40-59 year old | 2,555,547 | 0** | 87,673 | 19 | 33.56 | - |
| 60-74 year old | 1,608,239 | 9,835 | 166,642 | 144 | 19.17 | 3.97 |
| 75-84 year old | 654,675 | 23,003 | 98,102 | 433 | 9.68 | 2 |
| 85+ year old | 239,958 | 33,502 | 39,759 | 612 | 4.63 | 0.96 |

\* Distribution in age groups is imputed as described in Methods section
\*\* officially there are some individuals below the age of 60 in nursing homes. We decided to not include those as they very few

## RESULTS

### Descriptives for infections and other features

In 2024, a total of 1,212 COVID-19 deaths were reported in Austria.^28^ In our previous publication based on wastewater data, we estimated 2.5 million SARS-CoV-2 infections (Table 1). See Table 2 for the allocation of those infections and deaths to the individual strata.

**Table 2:** Allocated SARS-COV-2 infections and COVID-19 deaths per strata in Austria in 2024.

| Value | vaccination<br>status | nursing<br>home<br>residency | age range (years) |  |  |  |  |  |
| --- | --- | --- | --- | --- | --- | --- | --- | --- |
|  |  |  | <20 | 20-39 | 40-59 | 60-74 | 75-84 | 85+ |
| Infections |  |  |  |  |  |  |  |  |
|  | unvacc | yes | 0 | 0 | 0 | 2,455 | 5,498 | 7,883 |
|  | unvacc | no | 478,450 | 631,388 | 677,614 | 398,973 | 150,963 | 48,580 |
|  | vacc | yes | 0 | 0 | 0 | 227 | 775 | 1,252 |
|  | vacc | no | 1,902 | 8,881 | 19,258 | 36,896 | 21,287 | 7,718 |
| Deaths |  |  |  |  |  |  |  |  |
|  | unvacc | yes | 0 | 0 | 0 | 9 | 43 | 96 |
|  | unvacc | no | 1 | 3 | 19 | 124 | 344 | 443 |
|  | vacc | yes | 0 | 0 | 0 | 1 | 5 | 13 |
|  | vacc | no | 0 | 0 | 0 | 10 | 41 | 60 |

Note that we present rounded values, but use decimals for calculations. Total deaths per age group are publicly available, further stratification of deaths and all stratification of infections are however based on the allocation process described in the Methods section. Allocation is dependent on VEI, VED, age group infection probability (AIP) and nursing home infection probability (NIP) parameter choices. Presented allocation in this table is based on main analysis parameter values (VEI = 20%; VED = 15%; AIP and NIP equal across groups).
Vaccinated = vaccinated in 2024, unvaccinated = not vaccinated in 2024

### Infection fatality rate

Overall IFR in 2024 in Austria was 0.048%, with estimated IFR for unvaccinated and vaccinated being 0.045% and 0.13%, respectively (Table 3). Vaccinated had a higher estimated IFR as their average age was substantially higher (Table 1). IFR had a steep age gradient (Table 3). IFR of the oldest age group (85+ year old) was 0.91% and 0.77% for unvaccinated and vaccinated community dweller, respectively, and 1.22% and 1.04% for unvaccinated and vaccinated nursing home residents, respectively (Table 3). Their total IFR irrespective of nursing home residency was 0.95% for unvaccinated and 0.81% for the vaccinated.

**Table 3:** IFRs of all strata, based on age group, nursing home residency and vaccination in 2024.

|  | **community dweller IFR** |  | **nursing home resident IFR** |  | **total IFR** |  |
| --- | --- | --- | --- | --- | --- | --- |
|  | **unvaccinated** | **vaccinated** | **unvaccinated** | **vaccinated** | **unvaccinated** | **vaccinated** |
| Total | 0.039% | 0.116% | 0.94% | 0.84% | 0.045% | 0.13% |
| 0-19 | 0.00021% | 0.00018% | - | - | 0.00021% | 0.00018% |
| 20-39 | 0.00047% | 0.00040% | - | - | 0.00047% | 0.00040% |
| 40-59 | 0.0027% | 0.0023% | - | - | 0.0027% | 0.0023% |
| 60-74 | 0.031% | 0.027% | 0.36% | 0.31% | 0.033% | 0.028% |
| 75-84 | 0.23% | 0.19% | 0.78% | 0.66% | 0.25% | 0.21% |
| 85+ | 0.91% | 0.77% | 1.22% | 1.04% | 0.95% | 0.81% |

Note that totals show total may show higher IFR in vaccinated due to vaccinated being much older on average
Vaccinated = vaccinated in 2024, unvaccinated = not vaccinated in 2024

Nursing home residency had the largest impact in the relatively youngest group (60-74), in which community dwellers had IFRs of 0.031% (unvaccinated) and 0.027% (vaccinated) but nursing home residents had IFRs of 0.36% (unvaccinated) and 0.31% (vaccinated). Overall nursing home IFRs were 0.94% for unvaccinated and 0.84% for vaccinated (Table 3).

### Number needed to vaccinate to prevent one COVID-19 death and for one LYS

Estimated NNV (LYS) in the age groups 0-19, 20-39, 40-59, 60-74 and 75-84 years were very high. They were 5,497,526 (151,570), 2,432,498 (92,614), 415,714 (24,777), 35,925 (3,748), and 4,882 (1,009), respectively, in community dwellers. For nursing home residents, they were 3,105 (1,565) in 60-74 years and 1,423 (1,423) in 75-84 years.

For 2024, estimated NNV in 85+ year old community dwellers and nursing home residents was 1,215 and 907, respectively. Sensitivity range analyses results for NNV for 85+ year olds ranged from 770 to 2,065 for community dwellers and 575 to 1,652 in nursing home residents (Table 4). When accounting for all possible combinations of sensitivity range values, NNV in the 85+ years old ranged from 605 to 4,144 and 452 to 3,093 in community dwellers and nursing home residents, respectively (Table S2).

**Table 4:**
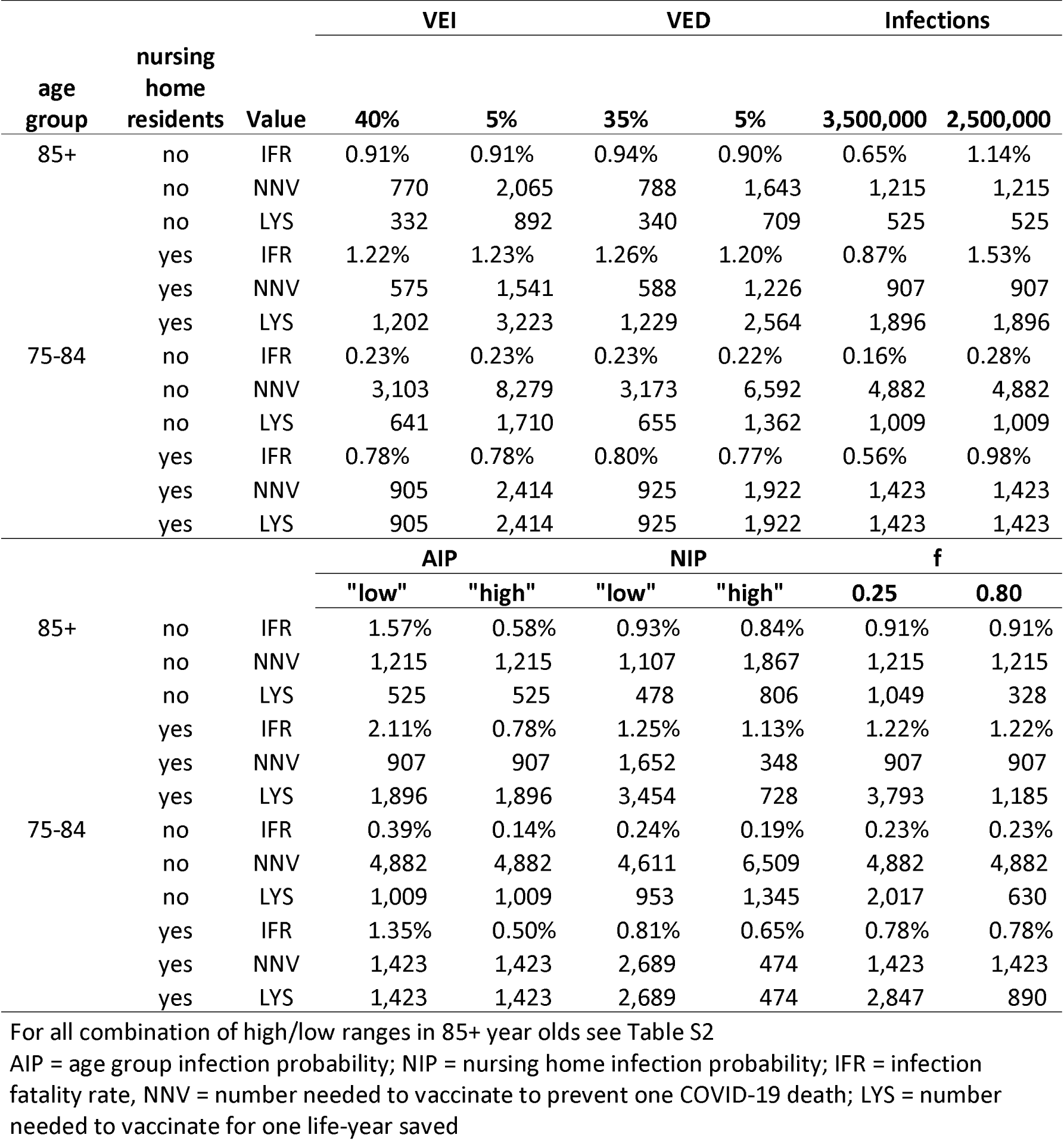
Sensitivity range estimates of IFR, NNV and LYS of unvaccinated 75-84 and 85+ age groups.

The estimated LYS for the 85+ year olds in 2024 in Austria was 525 for community dwellers and 1,896 for nursing home residents. Sensitivity analyses showed values in LYS of 85+ year olds ranging from 328 to 1,049 for community dwellers and from 728 to 3,793 for nursing home residents (Table 4). In all possible combinations of sensitivity ranges in 85+ year olds, the estimated LYS ranged from 163 to 3,578 for community dwellers and from 590 to 12,932 for nursing home residents (Table S2).

Table 4 shows the outcomes for different parameter sensitivity ranges in the two most relevant (oldest) age groups (75-84 and 85+ year olds). Besides the 85+ years age group, LYS <1,000 were seen also for the 75-84 years old group under favourable sensitivity assumptions in VEI, VED, NIP, and f. The best (lowest) LYS for the 75-84 years old group was 630 and 474 for community-dwelling and nursing home residents, respectively. For younger than 75 years old age groups, all sensitivity analyses show very high NNV and LYS (not shown).

Combinations between low and high parameter ranges to investigate the outmost outliers in our estimations appear in Table S2.

## DISCUSSION

We estimate that in 2024 in Austria the total IFR was at 0.048%. IFRs were below 1% for all age and nursing home stratified groups expect the 85+ year old nursing home residents. Estimated NNV were extremely high in young age groups (e.g., for 0-19 and 20-39 year old individuals, they were 5,497,526 and 2,432,498, respectively), and were very high even in older individuals. Even in the oldest age stratum (85+ years old) an estimated 1,215 doses would have been needed to avoid a single COVID-19 death in community dwelling individuals and 907 in nursing home residents. For one LYS, values exceeded 1,000 for all age strata both in community dwelling and nursing home residents, with the exception of 85+ year old community dwelling individuals. For those 75-84 years old, LYS <1,000 could be seen only with some of the most favourable sensitivity range assumptions, and no LYS values in this age group were lower than 474.

Our IFR estimates match the estimates towards the end of the official pandemic in May 2023.^4,5^ Thus, even though the number of administered vaccinations decreased between 2023 and 2024, the estimated fatality rate of COVID-19 did not change.^26^ This is likely generalisable to other western countries with comparable social and health care structures.

Importantly our results strongly argue against recommendations for COVID-19 booster vaccinations in children and healthy adults, as the NNV and LYS were extraordinary high for these groups (Table 4).^7,8^ Even for most older individuals, NNV and LYS are quite high and may lead to unfavourable cost-effectiveness with current prices of COVID-19 vaccines.

Our estimated NNV and LYS are lower than previous estimates for earlier timeframes, indicating a decrease in the potential benefit of COVID-19 vaccines.^35,39^ This may be partially due to a decrease in severity of COVID-19 infections, most likely driven by currently dominant high-immune escape SARS-CoV-2 variants and virtually universal levels of natural and/or hybrid immunisation in the population.^5^ In addition, we use slightly lower VE estimates than previous investigations, because the few existing recent estimates showed lower (if any) VE at later stages of the pandemic and post-pandemic.^12,15^ Importantly, recent studies have documented a healthy vaccinee effect, suggesting that previous estimates of VE may have been overestimations as vaccine recipients were healthier than those not vaccinted.^16–18^

We acknowledge several limitations of our calculations. We rely on some assumptions which may introduce bias into our analyses. This mainly concerns the parameter choices for allocation of infections and deaths. VEI and VED values specifically may be under- or overestimated. On one hand a recent case control study indicates a VE of 48% COVID-19– associated critical illness.^15^ On the other hand, publications on the healthy vaccinee effect suggest that the actual VE is likely lower.^16–18^ We aimed to address these concerns in the sensitivity analyses by varying multiple assumed parameters. The assumption that everyone was previously infected may also lead to an underestimation of IFR and VE, if previously non-infected individuals exist. However, it is generally agreed upon that almost all people (other than new-borns) have had a previous infection.^1^ As the youngest cohorts have a quasi-non-existent risk to die of a COVID-19 infection, those individuals do not affect the appropriateness of the assumption in the present study.^4^

Further, we did not account for quality-adjusted life years (QALY) and other potentially relevant outcomes (e.g., hospitalisations, post-acute sequalae) or rare but documented negative effects of vaccinations such as thrombosis after adenovirus vector-based vaccines or myocarditis after mRNA vaccines (more prevalent in young men).^40,41^ Additional investigations to consider multiple outcomes and studies on formal cost-effectiveness of COVID-19 vaccines are warranted. However, deaths are the most important outcome. A formal cost-effectiveness analysis for Canada in 2024-2025 estimated a favourable ICER of $21,227 per QALY for a program focusing on COVID-19 vaccination in those over 65 years old and younger people with medical conditions.^42^ However, this calculation assumed 40% of the list price (only $43 per dose). The incremental cost-effectiveness ratio (ICER) would thus be unfavourable with full vaccine price. Moreover, some of the assumptions made in that study and in some other pre-emptive cost-effectiveness assessments by some other vaccination advisory organizations have probably led to too optimistic cost-effectiveness expectations.^43,44^

Finally, we considered that there is no misclassification regarding causes of death listed as COVID-19. However, there is accumulating evidence that in the Omicron period, in a large share of the death certificates with COVID-19 death listed, the deaths were not in fact due to SARS-CoV2.^45,46^ If so, the IFR may be overestimated and NNV and LYS may be underestimated substantially. This would make vaccination and even less desirable policy even in very old individuals.

In conclusion, COVID-19 vaccinations were crucial throughout the pandemic, but, our 2024 estimates indicate that additional COVID-19 vaccinations for the general population are unlikely to offer significant benefit or to be cost-effective. An exception may be potentially the very oldest cohort, but even in that age group the cost-benefit can be questioned. These findings are crucial for the continued management of SARS-CoV-2 both within and beyond healthcare settings.

## Contributors

UR and SP conceptualized the study with inputs from JPAI. UR wrote the original draft of the manuscript. SP acquired funding. UR wrote software. UR performed the formal analyses. SP administered the project. All authors contributed to writing, reviewing, and editing the manuscript, and approved the final version before submission.

## Data sharing statement

All data that support the findings are publicly available.

## Data Availability

All data that support the findings are publicly available.

## Acknowledgements

The authors thank all persons and organizations involved in data collection.

