## Supplements for "Nationwide estimates of SARS-CoV-2 infection fatality rates and numbers needed to vaccinate for SARS-CoV-2 vaccines in 2024 in Austria"

**Table S1:** STROBE Statement—Checklist of items for *cohort studies*

| | Item No | Recommendation | Main text page |
|---|---|---|---|
| **Title and abstract** | 1 | (*a*) Indicate the study's design with a commonly used term in the title or the abstract | Page 2, Abstract |
| | | (*b*) Provide in the abstract an informative and balanced summary of what was done and what was found | Page 2 , Abstract |
| **Introduction** | | | |
| Background/rationale | 2 | Explain the scientific background and rationale for the investigation being reported | Page 4 and 5, Introduction |
| Objectives | 3 | State specific objectives, including any prespecified hypotheses | Page 4 and 5, Introduction |
| **Methods** | | | |
| Study design | 4 | Present key elements of study design early in the paper | Page 5, Methods |
| Setting | 5 | Describe the setting, locations, and relevant dates, including periods of recruitment, exposure, follow-up, and data collection | Page 5 to 7 , Methods |
| Participants | 6 | (*a*) Give the eligibility criteria, and the sources and methods of selection of participants. Describe methods of follow-up | Page 5 to 7, Methods |
| | | (*b*) For matched studies, give matching criteria and number of exposed and unexposed | Not applicable |
| Variables | 7 | Clearly define all outcomes, exposures, predictors, potential confounders, and effect modifiers. Give diagnostic criteria, if applicable | Page 3,4 and 7 to 9 , Introduction and Methods |
| Data sources/ measurement | 8* | For each variable of interest, give sources of data and details of methods of assessment (measurement). Describe comparability of assessment methods if there is more than one group | Page 4 and 7, Methods |
| Bias | 9 | Describe any efforts to address potential sources of bias | Page 5 to 9, Methods |
| Study size | 10 | Explain how the study size was arrived at | Not applicable; Nationwide study |
| Quantitative variables | 11 | Explain how quantitative variables were handled in the analyses. If applicable, describe which groupings were chosen and why | Page 5, 6 and 7, Methods |
| Statistical methods | 12 | (*a*) Describe all statistical methods, including those used to control for confounding | Not applicable |
| | | (*b*) Describe any methods used to examine subgroups and interactions | Page 3 to 9, Methods (and Supplements) |
| | | (*c*) Explain how missing data were addressed | Not applicable. |
| | | (*d*) If applicable, explain how loss to follow-up was addressed | Not applicable |
| | | (*e*) Describe any sensitivity analyses | Page 7 and 8, Methods |
| **Results** | | | |
| Participants | 13* | (a) Report numbers of individuals at each stage of study—eg numbers potentially eligible, examined for eligibility, | Page 9 and 10, Results: Table 1 and 2 |

| | | | |
|---|---|---|---|
| | | confirmed eligible, included in the study, completing follow-up, and analysed | |
| | | (b) Give reasons for non-participation at each stage | Not applicable |
| | | (c) Consider use of a flow diagram | |
| Descriptive data | 14* | (a) Give characteristics of study participants (eg demographic, clinical, social) and information on exposures and potential confounders | Page 9; Table 1 |
| | | (b) Indicate number of participants with missing data for each variable of interest | no missing data |
| | | (c) Summarise follow-up time (eg, average and total amount) | Not applicable |
| Outcome data | 15* | Report numbers of outcome events or summary measures over time | Page 9 to 13, Results; Tables and text |
| Main results | 16 | (*a*) Give unadjusted estimates and, if applicable, confounder-adjusted estimates and their precision (eg, 95% confidence interval). Make clear which confounders were adjusted for and why they were included | Not applicable |
| | | (*b*) Report category boundaries when continuous variables were categorized | Not applicable |
| | | (*c*) If relevant, consider translating estimates of relative risk into absolute risk for a meaningful time period | Not applicable |
| Other analyses | 17 | Report other analyses done—eg analyses of subgroups and interactions, and sensitivity analyses | Page 12 and 13 , Results (and Supplements; Table S2) |
| **Discussion** | | | |
| Key results | 18 | Summarise key results with reference to study objectives | Page 13, Discussion |
| Limitations | 19 | Discuss limitations of the study, taking into account sources of potential bias or imprecision. Discuss both direction and magnitude of any potential bias | Page 14 , Discussion |
| Interpretation | 20 | Give a cautious overall interpretation of results considering objectives, limitations, multiplicity of analyses, results from similar studies, and other relevant evidence | Page 13 and 14, Discussion |
| Generalisability | 21 | Discuss the generalisability (external validity) of the study results | Page 13 and 14, Discussion |
| **Other information** | | | |
| Funding | 22 | Give the source of funding and the role of the funders for the present study and, if applicable, for the original study on which the present article is based | Page 15 |

*Give information separately for exposed and unexposed groups.

**Note:** An Explanation and Elaboration article discusses each checklist item and gives methodological background and published examples of transparent reporting. The STROBE checklist is best used in conjunction with this article (freely available on the Web sites of PLoS Medicine at http://www.plosmedicine.org/, Annals of Internal Medicine at http://www.annals.org/, and Epidemiology at http://www.epidem.com/). Information on the STROBE Initiative is available at http://www.strobe-statement.org.

### Supplementary Methods

Infections and deaths are allocated to different strata [age group, vaccination 2024 (yes/no) and nursing home residency (yes/no)] which are used for calculation of IFR and consequently NNV and LYS as described in the main manuscript.

To calculate stratum specific IFRs we use published estimates of infection probabilities in different age groups in the US.[1] We also use infection probabilities for nursing home residents, based on early seroprevalence data of nurses in nursing homes and in local elderly care.[2] Infections are distributed to vaccinated and unvaccinated groups based on VEI estimates. Deaths distribution to vaccinated and unvaccinated groups (per age group) is based on infection distribution and VED estimates. Deaths (per age group and vaccination status) is allocated to nursing home residents or community dwellers based on distribution of deaths in the pandemic in Austria (Table 1).

The allocation of infections and deaths based on VEI and VED relies on the fact, that they represent conditional vaccine effectiveness estimates.[3] By definition, VEI quantifies how likely vaccinated individuals get infected (compared to unvaccinated individuals) and VED quantifies how likely those infected vaccinated individuals are to die (compared to infected unvaccinated individuals).

#### Distribution of infections to strata

Infections are allocated to age, nursing home residency (NHR), and vaccination status groups using population size and relative infection risk. Vaccination reduces the risk of infection by a factor equal to vaccine effectiveness against infection (VEI). Within each age group, the vaccinated population has an infection risk of 1 - VEI, while the unvaccinated has a risk of 1.

To allocate infections:

Adjust infection risk:

1. For each age group:

$$inf\_risk_{vacc} = V * (1 - VEI),$$

$$inf\_risk_{unvacc} = U$$

   where V and U are the number of vaccinated and unvaccinated individuals in the group.

2. Total risk and proportional allocation:

   The total risk is:

$$total_{risk} = inf\_risk_{vacc} + inf\_risk_{unvacc}$$

   infections are then distributed proportionally:

$$infected_{vacc} = infected_{total} * \frac{inf\_risk_{vacc}}{total_{risk}},$$

$$infected_{unvacc} = infected_{total} * \frac{inf\_risk_{unvacc}}{total_{risk}}$$

3. Adjustment by age-specific infection modifier:

   Infection allocation is weighted by age group infection probability (AIP) and group population size to account for possible differences in exposure or reporting between age groups. The AIP was set to 1 for all age groups (main analysis).[1]

4. Nursing home stratification:

   Within each vaccination group, infections are split between NHR and non-NHR based on their share in the total population of that age group as well as a nursing home infection probability (NIP). For the main analysis NIP was once again set to 1 for all age groups.

**Distribution of deaths to strata**

Allocation of deaths differs in two points: (1) distribution is calculated within age group (as death per age is available), and (2) distribution to nursing home residents is based on previously recorded mortality rates additionally to group size.[4]

Deaths are allocated within each age group based on the number of infections and the vaccine effectiveness against death (VED).

1. Death risk by vaccination status:

   For each group:

$$death\_risk_{vacc} = infected_{vacc} * (1 - VED),$$

$$death\_risk_{unvacc} = infected_{unvacc}$$

2. Proportional allocation of deaths:

   Total death risk:

$$death\_risk_{total} = death\_risk_{vacc} + death\_risk_{unvacc}$$

   Deaths are distributed proportionally:

$$deaths_{vacc} = D * \frac{deaths\_risk_{vacc}}{deaths\_risk_{total}},$$

$$deaths_{unvacc} = D * \frac{deaths\_risk_{unvacc}}{deaths\_risk_{total}}$$

   where D is the total number of deaths in the age group.

3. Nursing home adjustment:

Within each vaccination group, deaths are further divided between NHR and non-NHR using weights that reflect higher mortality risk in nursing homes. The adjustment is based on age-specific NHR mortality multipliers derived from a previously published study on COVID-19 mortality in Austria.[4]

4. Final Rounding and Correction

   After all allocations, infection and death counts are rounded while ensuring the overall totals remain correct. This is done by:
   - Flooring all counts
   - Calculating the remainder needed to reach the total
   - Adding 1 to the rows with the largest remainders until the correct total is restored

**Data preparation**
Vaccination data is available, but for different age groups than we are interested in.[5] Thus, we used population weighted estimates of single years in a group to re-allocate vaccinations to our age group distribution. I.e., we took e.g., the 30-44 year old category form the vaccination data and distributed the number of vaccinations in this group to every year in this range based on the population size of that age group (projected, mid 2024[6]). We were then able to add the 40-44 year old estimates to the available 45-59 year old data to get estimates of vaccinations in our 40-59 year old age group.

**Implicit Assumptions**
We use one value for VEI and VED. However there are clear indication of waning (especially in VEI) which we do not incorporate.[7] Thus the real VE would decrease with time from vaccination (Figure S1). By not accounting for this we implicitly assume an optimal vaccination time (right before the COVID-19 wave) for all vaccinations.[7] Note that yearly timing of SARS-CoV-2 waves is inconsistent,[8] thus in practice defining the optimal time for vaccination is challenging, and VE changes depending on the time since vaccination.

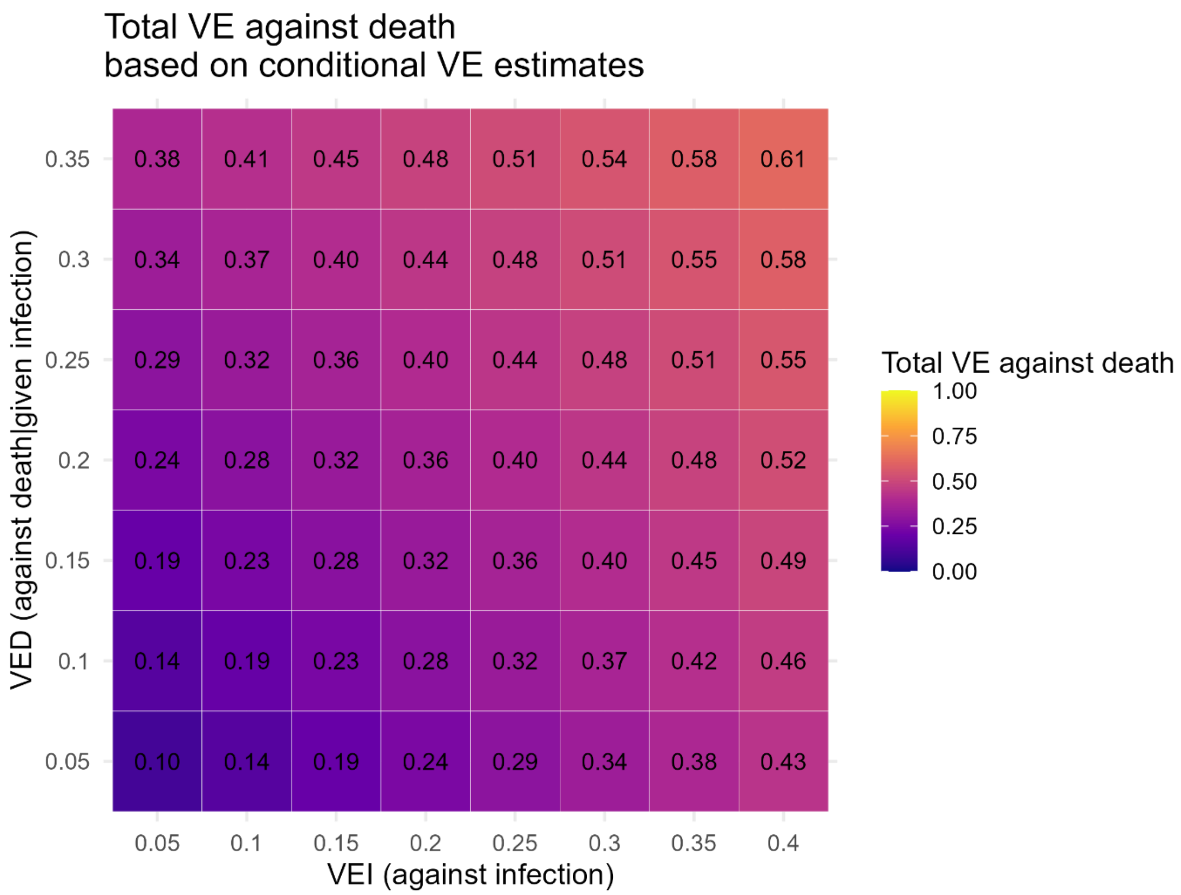

**Figure S1:** Plot of different VEI and VED combinations and their respective unconditional VE.

Note that due to the established estimate for the number of infections, a decrease in VEI increases the IFR estimates for the non-vaccinated group (given positive VED values) as relatively more infections are allocated to vaccinated group. This underlines the flexibility of this approach to estimate individual strata IFRs.

### Supplementary Results

**Table S2:** Sensitivity range estimates of IFR, NNV and LYS of unvaccinated 85+ year olds, for all possible combinations of low and high parameter estimates.

| nursing home residents | value | VEI=5%; VED=5%; Inf=2m; AIP=low; NIP = low; f = 0.25 | VEI=40%; VED=5%; Inf=2m; AIP=low; NIP = low; f = 0.25 | VEI=5%; VED=35%; Inf=2m; AIP=low; NIP = low; f = 0.25 | VEI=40%; VED=35%; Inf=2m; AIP=low; NIP = low; f = 0.25 | VEI=5%; VED=5%; Inf=3.5m; AIP=low; NIP = low; f = 0.25 | VEI=40%; VED=5%; Inf=3.5m; AIP=low; NIP = low; f = 0.25 | VEI=5%; VED=35%; Inf=3.5m; AIP=low; NIP = low; f = 0.25 | VEI=40%; VED=35%; Inf=3.5m; AIP=low; NIP = low; f = 0.25 |
|---|---|---|---|---|---|---|---|---|---|
| no | IFR | 1.94% | 1.94% | 2.04% | 2.00% | 1.11% | 1.11% | 1.17% | 1.14% |
| no | NNV | 4,144 | 887 | 1,006 | 605 | 4,144 | 887 | 1,006 | 605 |
| no | LYS | 3,578 | 766 | 868 | 523 | 3,578 | 766 | 868 | 523 |
| yes | IFR | 2.60% | 2.60% | 2.73% | 2.68% | 1.49% | 1.48% | 1.56% | 1.53% |
| yes | NNV | 3,093 | 662 | 751 | 452 | 3,093 | 662 | 751 | 452 |
| yes | LYS | 12,932 | 2,768 | 3,138 | 1,889 | 12,932 | 2,768 | 3,138 | 1,889 |
| nursing home residents | value | VEI=5%; VED=5%; Inf=2m; AIP=high; NIP = low; f = 0.25 | VEI=40%; VED=5%; Inf=2m; AIP=high; NIP = low; f = 0.25 | VEI=5%; VED=35%; Inf=2m; AIP=high; NIP = low; f = 0.25 | VEI=40%; VED=35%; Inf=2m; AIP=high; NIP = low; f = 0.25 | VEI=5%; VED=5%; Inf=3.5m; AIP=high; NIP = low; f = 0.25 | VEI=40%; VED=5%; Inf=3.5m; AIP=high; NIP = low; f = 0.25 | VEI=5%; VED=35%; Inf=3.5m; AIP=high; NIP = low; f = 0.25 | VEI=40%; VED=35%; Inf=3.5m; AIP=high; NIP = low; f = 0.25 |
| no | IFR | 0.72% | 0.71% | 0.75% | 0.74% | 0.41% | 0.41% | 0.43% | 0.42% |
| no | NNV | 4,144 | 887 | 1,006 | 605 | 4,144 | 887 | 1,006 | 605 |
| no | LYS | 3,578 | 766 | 868 | 523 | 3,578 | 766 | 868 | 523 |
| yes | IFR | 0.96% | 0.96% | 1.01% | 0.99% | 0.55% | 0.55% | 0.58% | 0.57% |
| yes | NNV | 3,093 | 662 | 751 | 452 | 3,093 | 662 | 751 | 452 |
| yes | LYS | 12,932 | 2,768 | 3,138 | 1,889 | 12,932 | 2,768 | 3,138 | 1,889 |

continued…

| nursing home residents | value | VEI=5%; VED=5%; Inf=2m; AIP=low; NIP = high; f = 0.25 | VEI=40%; VED=5%; Inf=2m; AIP=low; NIP = high; f = 0.25 | VEI=5%; VED=35%; Inf=2m; AIP=low; NIP = high; f = 0.25 | VEI=40%; VED=35%; Inf=2m; AIP=low; NIP = high; f = 0.25 | VEI=5%; VED=5%; Inf=3.5m; AIP=low; NIP = high; f = 0.25 | VEI=40%; VED=5%; Inf=3.5m; AIP=low; NIP = high; f = 0.25 | VEI=5%; VED=35%; Inf=3.5m; AIP=low; NIP = high; f = 0.25 | VEI=40%; VED=35%; Inf=3.5m; AIP=low; NIP = high; f = 0.25 |
|---|---|---|---|---|---|---|---|---|---|
| no | IFR | 1.94% | 1.94% | 2.04% | 2.00% | 1.11% | 1.11% | 1.17% | 1.14% |
| no | NNV | 4,144 | 887 | 1,006 | 605 | 4,144 | 887 | 1,006 | 605 |
| no | LYS | 3,578 | 766 | 868 | 523 | 3,578 | 766 | 868 | 523 |
| yes | IFR | 2.60% | 2.60% | 2.73% | 2.68% | 1.49% | 1.48% | 1.56% | 1.53% |
| yes | NNV | 3,093 | 662 | 751 | 452 | 3,093 | 662 | 751 | 452 |
| yes | LYS | 12,932 | 2,768 | 3,138 | 1,889 | 12,932 | 2,768 | 3,138 | 1,889 |

| nursing home residents | value | VEI=5%; VED=5%; Inf=2m; AIP=high; NIP = high; f = 0.25 | VEI=40%; VED=5%; Inf=2m; AIP=high; NIP = high; f = 0.25 | VEI=5%; VED=35%; Inf=2m; AIP=high; NIP = high; f = 0.25 | VEI=40%; VED=35%; Inf=2m; AIP=high; NIP = high; f = 0.25 | VEI=5%; VED=5%; Inf=3.5m; AIP=high; NIP = high; f = 0.25 | VEI=40%; VED=5%; Inf=3.5m; AIP=high; NIP = high; f = 0.25 | VEI=5%; VED=35%; Inf=3.5m; AIP=high; NIP = high; f = 0.25 | VEI=40%; VED=35%; Inf=3.5m; AIP=high; NIP = high; f = 0.25 |
|---|---|---|---|---|---|---|---|---|---|
| no | IFR | 0.72% | 0.71% | 0.75% | 0.74% | 0.41% | 0.41% | 0.43% | 0.42% |
| no | NNV | 4,144 | 887 | 1,006 | 605 | 4,144 | 887 | 1,006 | 605 |
| no | LYS | 3,578 | 766 | 868 | 523 | 3,578 | 766 | 868 | 523 |
| yes | IFR | 0.96% | 0.96% | 1.01% | 0.99% | 0.55% | 0.55% | 0.58% | 0.57% |
| yes | NNV | 3,093 | 662 | 751 | 452 | 3,093 | 662 | 751 | 452 |
| yes | LYS | 12,932 | 2,768 | 3,138 | 1,889 | 12,932 | 2,768 | 3,138 | 1,889 |

continued…

| nursing home residents | value | VEI=5%; VED=5%; Inf=2m; AIP=low; NIP = low; f = 0.8 | VEI=40%; VED=5%; Inf=2m; AIP=low; NIP = low; f = 0.8 | VEI=5%; VED=35%; Inf=2m; AIP=low; NIP = low; f = 0.8 | VEI=40%; VED=35%; Inf=2m; AIP=low; NIP = low; f = 0.8 | VEI=5%; VED=5%; Inf=3.5m; AIP=low; NIP = low; f = 0.8 | VEI=40%; VED=5%; Inf=3.5m; AIP=low; NIP = low; f = 0.8 | VEI=5%; VED=35%; Inf=3.5m; AIP=low; NIP = low; f = 0.8 | VEI=40%; VED=35%; Inf=3.5m; AIP=low; NIP = low; f = 0.8 |
|---|---|---|---|---|---|---|---|---|---|
| no | IFR | 1.94% | 1.94% | 2.04% | 2.00% | 1.11% | 1.11% | 1.17% | 1.14% |
| no | NNV | 4,144 | 887 | 1,006 | 605 | 4,144 | 887 | 1,006 | 605 |
| no | LYS | 1,118 | 239 | 271 | 163 | 1,118 | 239 | 271 | 163 |
| yes | IFR | 2.60% | 2.60% | 2.73% | 2.68% | 1.49% | 1.48% | 1.56% | 1.53% |
| yes | NNV | 3,093 | 662 | 751 | 452 | 3,093 | 662 | 751 | 452 |
| yes | LYS | 4,041 | 865 | 981 | 590 | 4,041 | 865 | 981 | 590 |

| nursing home residents | value | VEI=5%; VED=5%; Inf=2m; AIP=high; NIP = low; f = 0.8 | VEI=40%; VED=5%; Inf=2m; AIP=high; NIP = low; f = 0.8 | VEI=5%; VED=35%; Inf=2m; AIP=high; NIP = low; f = 0.8 | VEI=40%; VED=35%; Inf=2m; AIP=high; NIP = low; f = 0.8 | VEI=5%; VED=5%; Inf=3.5m; AIP=high; NIP = low; f = 0.8 | VEI=40%; VED=5%; Inf=3.5m; AIP=high; NIP = low; f = 0.8 | VEI=5%; VED=35%; Inf=3.5m; AIP=high; NIP = low; f = 0.8 | VEI=40%; VED=35%; Inf=3.5m; AIP=high; NIP = low; f = 0.8 |
|---|---|---|---|---|---|---|---|---|---|
| no | IFR | 0.72% | 0.71% | 0.75% | 0.74% | 0.41% | 0.41% | 0.43% | 0.42% |
| no | NNV | 4,144 | 887 | 1,006 | 605 | 4,144 | 887 | 1,006 | 605 |
| no | LYS | 1,118 | 239 | 271 | 163 | 1,118 | 239 | 271 | 163 |
| yes | IFR | 0.96% | 0.96% | 1.01% | 0.99% | 0.55% | 0.55% | 0.58% | 0.57% |
| yes | NNV | 3,093 | 662 | 751 | 452 | 3,093 | 662 | 751 | 452 |
| yes | LYS | 4,041 | 865 | 981 | 590 | 4,041 | 865 | 981 | 590 |

continued…

| nursing home residents | value | VEI=5%; VED=5%; Inf=2m; AIP=low; NIP = high; f = 0.8 | VEI=40%; VED=5%; Inf=2m; AIP=low; NIP = high; f = 0.8 | VEI=5%; VED=35%; Inf=2m; AIP=low; NIP = high; f = 0.8 | VEI=40%; VED=35%; Inf=2m; AIP=low; NIP = high; f = 0.8 | VEI=5%; VED=5%; Inf=3.5m; AIP=low; NIP = high; f = 0.8 | VEI=40%; VED=5%; Inf=3.5m; AIP=low; NIP = high; f = 0.8 | VEI=5%; VED=35%; Inf=3.5m; AIP=low; NIP = high; f = 0.8 | VEI=40%; VED=35%; Inf=3.5m; AIP=low; NIP = high; f = 0.8 |
|---|---|---|---|---|---|---|---|---|---|
| no | IFR | 1.94% | 1.94% | 2.04% | 2.00% | 1.11% | 1.11% | 1.17% | 1.14% |
| no | NNV | 4,144 | 887 | 1,006 | 605 | 4,144 | 887 | 1,006 | 605 |
| no | LYS | 1,118 | 239 | 271 | 163 | 1,118 | 239 | 271 | 163 |
| yes | IFR | 2.60% | 2.60% | 2.73% | 2.68% | 1.49% | 1.48% | 1.56% | 1.53% |
| yes | NNV | 3,093 | 662 | 751 | 452 | 3,093 | 662 | 751 | 452 |
| yes | LYS | 4,041 | 865 | 981 | 590 | 4,041 | 865 | 981 | 590 |

| nursing home residents | value | VEI=5%; VED=5%; Inf=2m; AIP=high; NIP = high; f = 0.8 | VEI=40%; VED=5%; Inf=2m; AIP=high; NIP = high; f = 0.8 | VEI=5%; VED=35%; Inf=2m; AIP=high; NIP = high; f = 0.8 | VEI=40%; VED=35%; Inf=2m; AIP=high; NIP = high; f = 0.8 | VEI=5%; VED=5%; Inf=3.5m; AIP=high; NIP = high; f = 0.8 | VEI=40%; VED=5%; Inf=3.5m; AIP=high; NIP = high; f = 0.8 | VEI=5%; VED=35%; Inf=3.5m; AIP=high; NIP = high; f = 0.8 | VEI=40%; VED=35%; Inf=3.5m; AIP=high; NIP = high; f = 0.8 |
|---|---|---|---|---|---|---|---|---|---|
| no | IFR | 0.72% | 0.71% | 0.75% | 0.74% | 0.41% | 0.41% | 0.43% | 0.42% |
| no | NNV | 4,144 | 887 | 1,006 | 605 | 4,144 | 887 | 1,006 | 605 |
| no | LYS | 1,118 | 239 | 271 | 163 | 1,118 | 239 | 271 | 163 |
| yes | IFR | 0.96% | 0.96% | 1.01% | 0.99% | 0.55% | 0.55% | 0.58% | 0.57% |
| yes | NNV | 3,093 | 662 | 751 | 452 | 3,093 | 662 | 751 | 452 |
| yes | LYS | 4,041 | 865 | 981 | 590 | 4,041 | 865 | 981 | 590 |

AIP = age group infection probability; NIP = nursing home infection probability
